# Incivility in the workplace: A study of nursing and midwifery staff in Northern Ghana

**DOI:** 10.1101/2022.12.14.22283468

**Authors:** Alipitio Boo, Veronica Millicent Dzomeku, Abigail Kusi-Amponsah Diji, Adwoa Bemah Bonsu, Felix Apiribu

## Abstract

**Introduction:** The purpose of this study was to assess the prevalence, sources and effect of incivility in a tertiary hospital in Northern Ghana.

**Methods:** A descriptive cross-sectional survey was conducted among 321 nurses at a tertiary-level hospital in Northern Ghana. Between October and November 2022, participating nurses responded to an online or self-administered questionnaire using the 43-item incivility scale developed by Guidroz and colleagues. Data collected was cleaned in Microsoft Excel and transported into SPSS version 21 for further statistical analysis.

**Results:** Two-fifths of the participants (n=131, 40.8%) were aged 31-35 years. Over 50% of the participants were males (n=161), married (n=187) and had a Bachelor’s degree (n=166). A little over 30% of the participants had worked in the nursing profession for more than 8 years.

**Conclusion:** The participants reported an average incidence of 54.5% of incivility sometimes, most of the time, and all the time. Moreover, in terms of incivility reported under the subscales, the average greatest prevalence recorded was displaced frustration under patients/relations, with a rate of 78.9%. The least average prevalence rate indicated under the subscales was from abusive supervision under direct supervisors with a rate of 30.7%.

The existence of incivility in healthcare settings does not support a setting where healthcare workers and patients may operate safely. Therefore, it is advised that frequent in-service training sessions on what constitutes incivility is held for nurses and the hospital’s general staff in order to raise awareness of the negative impacts of incivility.

## Background

The nursing profession is rated highest based on Gallup polls on honesty/ethics globally (1). However, low intensity disruptive behaviours such as blaming, lying, gossiping, withholding important information, using abusive language, and verbal or physical intimidation exist in reality particularly in healthcare settings (2). The American Nurses Association refers to these behaviours as incivility. Incivility encompasses behaviours such as spreading rumours, gossiping, violent outbursts, yelling, condescending and the like in healthcare environments (2).

Incivility is very common in workplaces but more endemic in healthcare settings especially among nurses (3, 4). In industrialized nations such as the United States of America, the United Kingdom, and Europe, the prevalence of incivility among nurses is reported to be between 60% and 70% (5). Conner Black (5) argued that this prevalence rate could be more because the victims usually fear to report acts of incivility for fear of revenge and loss of job since the perpetrators are mostly in influential positions. Another reason was the inability of healthcare settings to formulate policies to address the phenomenon of low intensity disruptive behaviours (5).

According to Hopkinson and colleagues (6), incivility among nurses in healthcare settings cost the United States of America more than $4 billion dollars annually due to increased staff turnover and nurses quitting their jobs as a result of uncivil behaviours from colleagues, physicians, supervisors and patients/family. Evidence suggests that incivility contributes to the root cause of ineffective teamwork and poor communication in healthcare settings (7). For instance, uncivil behaviours such as lack of respect for one another at the workplace, shouting, raising one’s voice, threats etc. by employees in healthcare settings result in ineffective collaboration and poor communication among team members leading to unsafe patient care (10, 11).

In the Middle East and Africa, ineffective teamwork and poor communication among healthcare professionals in healthcare settings account for about 83% of all preventable medical errors and 30% of all deaths (8). As reported by Houck and Colbert (7), the presence of incivility in healthcare settings promotes ineffective teamwork and poor communication among healthcare personnel.

There is a dearth of literature in Ghana and Africa that discusses the prevalence, sources and effect of incivility on nursing and patient outcomes in the healthcare environment. As a result, this study filled a knowledge vacuum in the body of literature.

## Methodology

### Study Design

A descriptive cross-sectional survey was deemed appropriate as the researchers wanted to assess the effect of incivility among nurses at a single point in time (9).

### Study Setting

The study was conducted at a tertiary-level hospital in Northern Ghana. The hospital has a total bed capacity of 800 and serves as a referral center for the northern part of the country and other neighboring countries. Outpatient and inpatient services are provided at the respective departments of the hospital (e.g., Polyclinic, Accident and Emergency, Paediatrics, General Surgery).

### Population, Sample and Sampling Technique

The study population comprised all cadre of nurses working in the hospital. At the time of data collection, the hospital had a total of 1711 nurses (1002 rendering inpatient services and 698 rendering outpatient services respectively). Nurses who had worked for a minimum period of one year were eligible for inclusion in the current study as this period was considered enough for reporting acts of incivility (12). Eligible nurses who were on any type of leave were excluded from participation.

The sample size was determined using the Krejcie-Morgan sample size determination formula, s = X^2NP(1-P)/d^2(N-1)+X^2(1-P) where s=sample size, X^2=the table value of chi-square for 1 degree of freedom at the desired confidence level (3.841), N=the population size=1711, P=the population proportion (assumed to be 50% since this would give the maximum sample size, d=the degree of accuracy expressed as a proportion (0.05). The sample size, s=3.841^2*1711*0.50(1-0.50)/0.05^2(1711-1) +3.841^2(1-0.50) =314. Therefore, the minimum sample size that would be recruited for this study was 314. However, a sample size of 321 respondents was used. Out of this number, participants were proportionally drawn (190 from inpatient and 131 from outpatient departments respectively). The hospital renders both primary(polyclinic) and tertiary healthcare services. As a result, nurses render services on both inpatient and outpatient bases. Therefore, participants were proportionally drawn.

### Inclusion and Exclusion Criteria

Nurses rendering inpatient and outpatient services at the hospital and willing to consent to participate in the study were included. Also, nurses on full time employment and who have worked for at least one year were included in the study.

However, nurses rendering inpatient and outpatient services at the hospital but were not at post at the time of data collection were excluded from the study. Nurses who also refuse to consent to participate in the study were not included in the study.

### Data Collection Procedure

At the outpatient department (polyclinic), the principal investigator seeks permission from the deputy director of nursing services to engage nurses on shift bases to outline the aim and objectives of the study to them for voluntary participation. Those who agreed to take part in the study telephone numbers or email addresses were recorded without their names to ensure anonymity and told they would be contacted at a later date for them to respond to a questionnaire at their convenient time via WhatsApp or email in google document format or printed questionnaires. Again, the same procedure was repeated at the inpatient department.

In all 217 participants responded to an online questionnaire using the nursing incivility scale while 104 participants responded to self-administered questionnaires using the same incivility scale. The data was collected between October and November, 2022.

Permission was first sought and received from the developers to use the nursing incivility scale via email communication. Also, permission was sought and received to adapt some questionnaires on the survey tool on Surveys on Patient Culture through an email communication too (https://www.ahrq.gov/sops/index.html). The nursing incivility scale tool is designed to assess nurses’ perceptions of incivility encountered in healthcare settings.

The reliability of the tool was evaluated using Cronbach’s alpha coefficient statistical method and the results showed that all of the values were much higher than the necessary minimum of 0.70 (13). The values varied from 0.81 to 0.94.

In addition, the content validity of the scale was evaluated by seven (7) experts, three of whom hold doctor of philosophy (PhD) in nursing and midwifery, one PhD candidate and three masters’ students with an overall content validity index of 0.87. Based on the scores and comments that emanated from the assessment, the statements on the incivility scale were modified to reflect frequency of occurrence of the phenomenon such as “not at all, few of the times, sometimes, most of the times, and all the time”.

### Data Collection Tool

The tool for data collection was a structured questionnaire and the nursing incivility scale. The questionnaires were used to collect data on participants’ demographics while the nursing incivility scale instrument was used to measure the prevalence, sources and effect of incivility among study participants. So, in all the data collection tool comprised four sections.

### Validity and Reliability

A pre-test of the questionnaires and the nursing incivility scale instrument was carried out among 50 nurses at secondary level hospital. The hospital is also located within the same metropolis where the research was conducted. As such their responses to the questionnaire items on the nursing incivility scale enabled the researchers modify if necessary to enhance clarity and precision. Validated questionnaires were then administered to the study respondents.

## Ethical Consideration

Ethical approval was obtained from the ethics committee (CHRPE/AP/456/22) of Kwame Nkrumah University of Science and Technology. In addition, a certificate of authorisation was obtained from the research department of the hospital prior to data collection. All data collected were treated confidential.

## Data Analysis

Data collected was cleaned in Microsoft Excel and transported into SPSS version 21 for further statistical analysis. Descriptive findings were stated in numbers and percentage distributions. Spearman Rank Correlations test (significant test) was performed to determine whether there was a correlation between the experiences of incivility by participants and patients and nursing outcomes. The patient and nursing outcomes measure of interest were safe work environment and intention to stay. The level of significance was accepted at alpha=0.05.

## Results

From table 1.1, the majority of the participants (n=131, 40.8%) were in the age range of 31-35. There was nearly equal distribution in terms of participants’ gender (n=161, 50.2% were males and n=160, 49.2% were females). In terms of marital status of participants, the majority were married (n=187, 58.3%). On participants’ level of education, the majority had obtained a first degree (n=166, 51.7%). In addition, most of the participants (n=155, 48.3%) respondents were working in the general inpatient department. Again, the majority of the participants (n=101, 31.5%) responded they have more than 8years of work experience. Furthermore, the majority of the participants (n=257, 80.1%) provide direct patient care and majority of the participants (n=246, 76.6%) run night shift three times in a week.

**Table 1.1.**
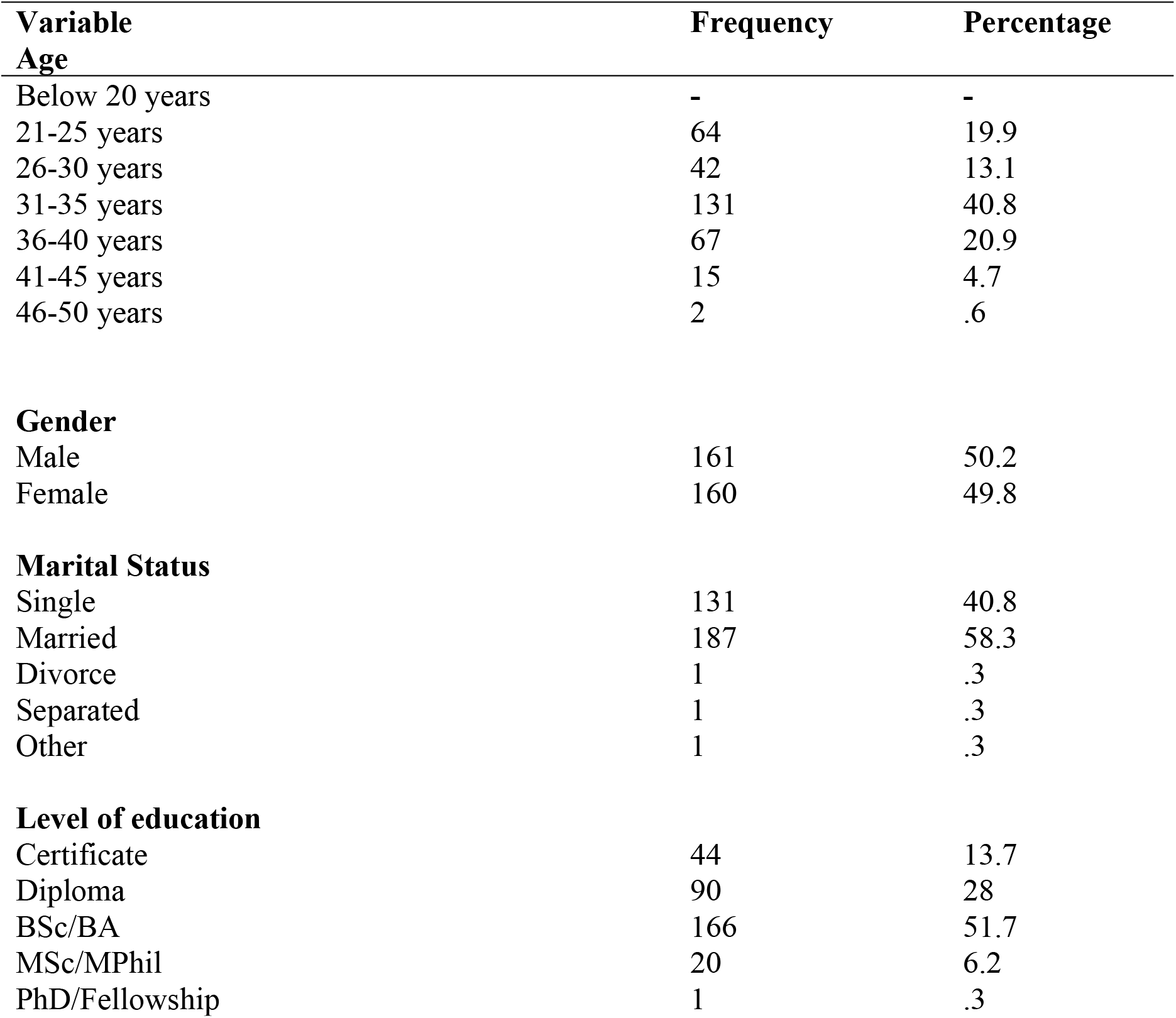
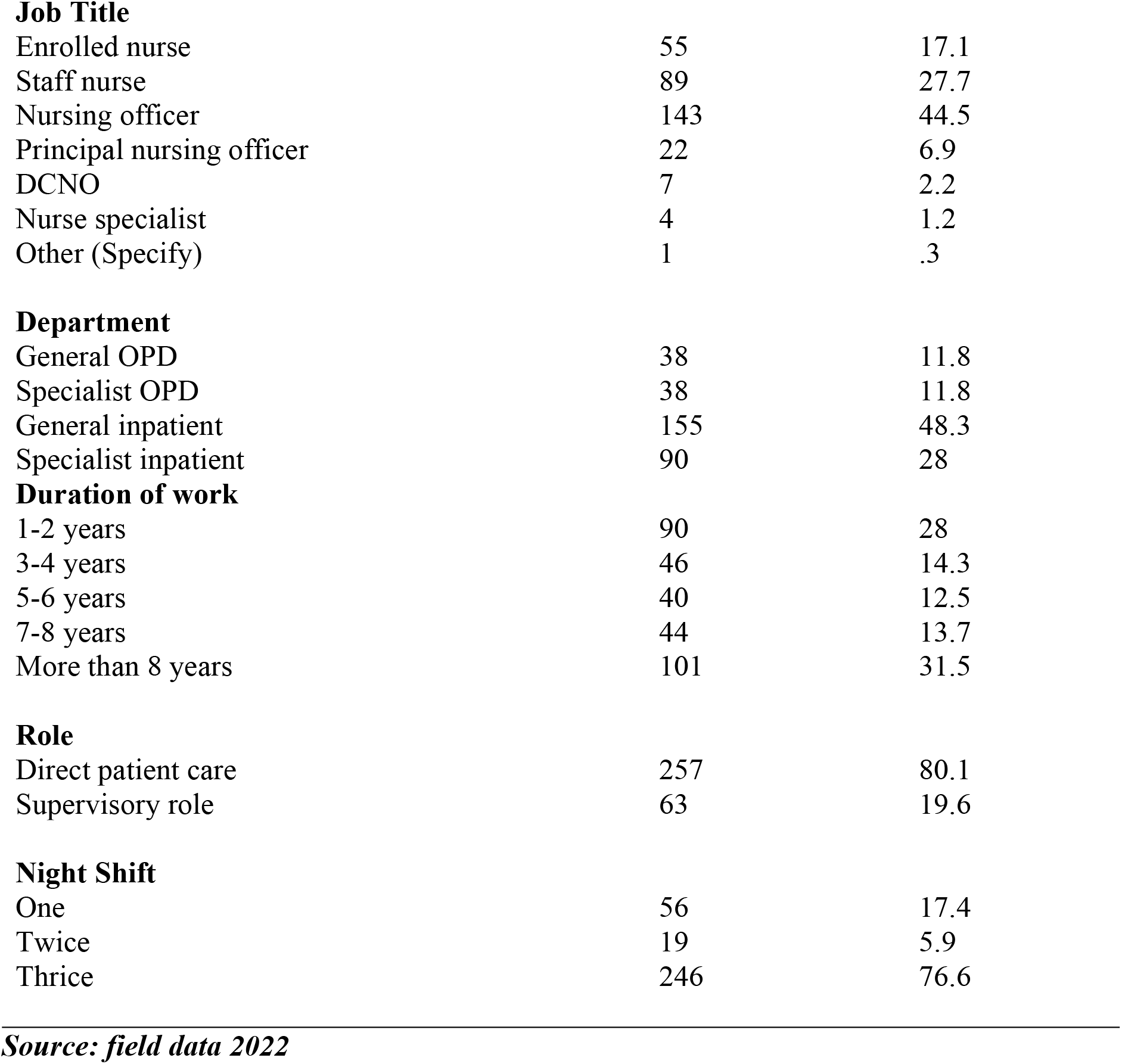
Participants’ demographic characteristics.

| **Variable** | **Frequency** | **Percentage** |
| --- | --- | --- |
| **Age** |  |  |
| Below 20 years | - | - |
| 21-25 years | 64 | 19.9 |
| 26-30 years | 42 | 13.1 |
| 31-35 years | 131 | 40.8 |
| 36-40 years | 67 | 20.9 |
| 41-45 years | 15 | 4.7 |
| 46-50 years | 2 | .6 |
| **Gender** |  |  |
| Male | 161 | 50.2 |
| Female | 160 | 49.8 |
| **Marital Status** |  |  |
| Single | 131 | 40.8 |
| Married | 187 | 58.3 |
| Divorce | 1 | .3 |
| Separated | 1 | .3 |
| Other | 1 | .3 |
| **Level of education** |  |  |
| Certificate | 44 | 13.7 |
| Diploma | 90 | 28 |
| BSc/BA | 166 | 51.7 |
| MSc/MPhil | 20 | 6.2 |
| PhD/Fellowship | 1 | .3 |
| **Job Title** |  |  |
| Enrolled nurse | 55 | 17.1 |
| Staff nurse | 89 | 27.7 |
| Nursing officer | 143 | 44.5 |
| Principal nursing officer | 22 | 6.9 |
| DCNO | 7 | 2.2 |
| Nurse specialist | 4 | 1.2 |
| Other (Specify) | 1 | .3 |
| **Department** |  |  |
| General OPD | 38 | 11.8 |
| Specialist OPD | 38 | 11.8 |
| General inpatient | 155 | 48.3 |
| Specialist inpatient | 90 | 28 |
| **Duration of work** |  |  |
| 1-2 years | 90 | 28 |
| 3-4 years | 46 | 14.3 |
| 5-6 years | 40 | 12.5 |
| 7-8 years | 44 | 13.7 |
| More than 8 years | 101 | 31.5 |
| **Role** |  |  |
| Direct patient care | 257 | 80.1 |
| Supervisory role | 63 | 19.6 |
| **Night Shift** |  |  |
| One | 56 | 17.4 |
| Twice | 19 | 5.9 |
| Thrice | 246 | 76.6 |

***Source: field data 2022***

### Prevalence and sources of incivility experienced by the participants

The prevalence of incivility experienced by respondents in the hospital settings (sometimes, most of the times, and all the time) are presented in the tables below.

From table 1.2, the overall average incivility rate reported by the participants’ experiences with incivility sometimes, most of the times, and all the time (frequently) under the nursing incivility scale was 54.5%. However, the highest average prevalence of incivility was reported from patients/relations with a rate of 65.2%. Additionally, in terms of incivility reported under the subscales, the average highest prevalence was from displaced frustration under patient/relation incivility with a rate of 78.9%.

**Table 1.1:** Prevalence of incivility experienced by the study participants.

| **Variable** | **Frequency (%)** |  |
| --- | --- | --- |
| **General invisibility** | **Rarely** | **Frequently** |
| 1. How often do hospital employees raise their voices when they get frustrated? | 81(25.2) | 240(74.7) |
| 2. How often do people blame others for their mistakes? | 83(25.9) | 238(74.1) |
| 3. How often do basic arguments turn into personal verbal attacks on other employees? | 109(33.9) | 212(66.1) |
| **Hostile Climate** | **28.3** | **71.6** |
| 4. How often do people make jokes about ethnic groups? | 121(37.7) | 200(62.3) |
| 5. How often do people make jokes about religious groups? | 157(48.9) | 164(51.1) |
| 6. How often do employees make inappropriate remarks about one's gender? | 183(57) | 138(43) |
| **Inappropriate Joke** | **47.9** | **52.1** |
| 7. How often do people take things without asking? | 104(32.4) | 217(67.6) |
| 8. How often do employees don't stick to an appropriate noise level (e.g., talking too loudly)? | 101(31.5) | 220(68.5) |
| 9. How often do employees display offensive body language (e.g., crossed arms, body posture)? | 138(43) | 183(57) |
| **Inconsiderate Behaviour** | **35.6** | **64.4** |
| **Grand prevalence from general (%)** | **37.3** | **62.7** |

| Variable | Frequency (%) |  |
| --- | --- | --- |
| **Incivility from Co-workers (Nurses)** | **Rarely** | **Frequently** |
| 1. How often do other nurses in your unit argue with each other frequently? | 158(49.2) | 163(50.8) |
| 2. How often do other nurses in your unit have heated arguments in the workplace? | 196(61.1) | 125(38.9) |
| 3. How often do other nurses in your unit have violent outbursts in the workplace? | 220(68.5) | 101(31.5) |
| **Hostile Climate** | **59.6** | **40.4** |
| 4. How often do other nurses in your unit scream at other employees? | 112(34.9) | 209(65.1) |
| 5. How often do other nurses in your unit gossip about one another? | 91(28.3) | 230(71.7) |
| 6. How often do other nurses in your unit gossip about their supervisor at work? | 94(29.3) | 227(70.7) |
| 7. How often do other nurses in your unit bad-mouth others in the workplace? | 120(37.4) | 201(62.6) |
| **Rumours/Gossip (%)** | **32.4** | **67.5** |
| 8. How often do other nurses in your unit spread bad rumours around here? | 123(38.3) | 198(61.7) |
| 9. How often do other nurses in your unit make little contribution to a project but expect to receive credit for working on it? | 144(44.9) | 177(55.1) |
| 10. How often do other nurses in your unit take credit for work they did not do? | 157(48.9) | 164(51.1) |
| **Free Riding (%)** | **44.0** | **56.0** |
| **Grand prevalence from co-workers (%)** | **45.4** | **54.6** |
| **Incivility from Direct Supervisors** | **Rarely** | **Frequently** |
| 1. My direct supervisor is verbally abusive | 237(73.8) | 84(26.2) |
| 2. My direct supervisor yells at me about matters that are not important. | 230(71.7) | 91(28.3) |
| 3. My direct supervisor shouted at me for making mistakes. | 211(65.7) | 110(34.3) |
| 4. My direct supervisor takes his/her feelings out on me (e.g., stress, anger, „blowing off steam) | 212(66.0) | 109(34.0) |
| **Abusive Supervision (%)** | **69.3** | **30.7** |
| 5. My direct supervisor does not respond to my concerns in a timely manner | 180(56.1) | 141(43.9) |
| 6. My direct supervisor demonstrates that she/he is superior to me | 133(41.4) | 188(58.6) |
| 7. My direct supervisor factors gossip into personnel decisions. | 177(55.1) | 144(44.9) |
| **Lack of respect (%)** | **50.9** | **49.1** |
| **Grand prevalence from direct supervisors** | **60.1** | **39.9** |
|  | Rarely | Frequently |
| **Incivility from Doctors/Physicians** |  |  |
| 1. To what extent are physicians verbally abusive? | 116(36.1) | 205(63.9) |
| 2. How often do physicians yell at you about matters that are not important? | 154(48.0) | 167(52) |
| 3. How often do physicians yell at you for making mistakes? | 183(57.0) | 138(43.0) |
| 4. How often do physicians take their feelings out on you (e.g., stress, anger, blowing off steam)? | 207(64.5) | 114(35.5) |
| **Abusive supervision (%)** | **51.4** | **48.6** |
| 5. How often do physicians not respond to your concerns in a timely manner? | 155(48.3) | 166(51.7) |
| 6. How often are you treated as though your time is not important? | 163(50.8) | 158(49.2) |
| 7. How often do physicians demonstrate superiority to you? | 147(45.8) | 174(54.2) |
| **Lack of respect (%)** | **48.3** | **51.7** |
| **Grand prevalence from physician/doctor (%)** | **49.8** | **50.2** |

| **Variable** | **Frequency (%)** |  |
| --- | --- | --- |
| **Incivility from Patients/Relations** | **Rarely** | **Frequently** |
| 1. How often do patients/visitors not trust the information you give them and ask to speak with someone of higher authority? | 214(66.7) | 107(33.3) |
| 2. How often do patients/visitors demonstrate superiority to nurses | 157(48.9) | 164(51.1) |
| 3. How often do patients/visitors make comments that question the competence of nurses? | 107(33.3) | 214(66.7) |
| 4. How often do patients/visitors criticise your job performance? | 192(59.8) | 129(40.2) |
| 5. How often do patients/visitors make personal verbal attacks against you? | 164(51.1) | 157(48.9) |
| 6. How often do patients/visitors pose unreasonable demands? | 102(31.8) | 219(68.2) |
| **Lack of respect (%)** | **48.6** | **51.4** |
| 7. How often do patients/visitors take out their frustrations on nurses? | 50(15.6) | 271(84.4) |
| 8. How often do patients/visitors make insulting comments to nurses? | 49(15.3) | 272(84.7) |
| 9. How often do patients/visitors treat nurses as if they were inferior? | 113(35.2) | 208(64.8) |
| 10. How often do patients/visitors show that they are impatient? | 58(18.1) | 263(81.9) |
| **Displaced Frustration (%)** | **21.1** | **78.9** |
| **Grand prevalence from patients/relations (%)** | **34.8** | **65.2** |

**Source: Field data 2022**

In conclusion, though the highest average percentage prevalence of incivilities were reported from general and patients/relations by the participants, the prevalence rate varied under the incivility subscales for general, co-workers, direct supervisors, doctors/physicians, and patients/relations.

### Effect of incivility on nursing and patient outcomes (safe work environment and intention to stay)

A Spearman Rank Correlations test was performed to determine whether there was a correlation between the experiences of incivility by participants and patients and nursing outcomes. In addition, an ANOVA test was performed to determine whether there was any statistical significance with the years of work experience of respondents and the experience of incivility from the various sources (general, co-worker, direct supervisors, doctors/physicians, and patients/relations). The results are presented in the tables below.

*** All p-values are highlighted and significant at alpha = 0.05

**Table 1.3:**
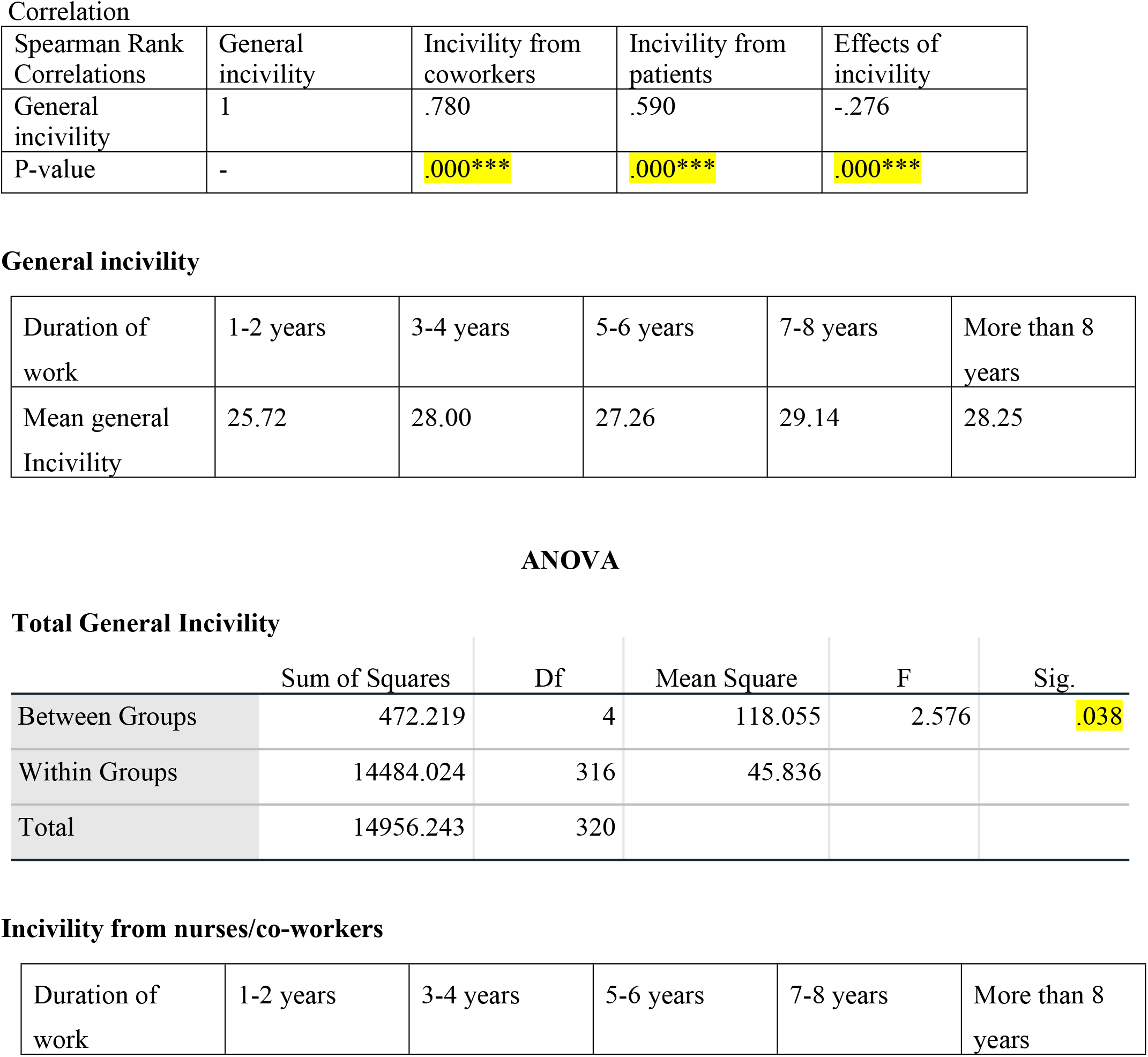
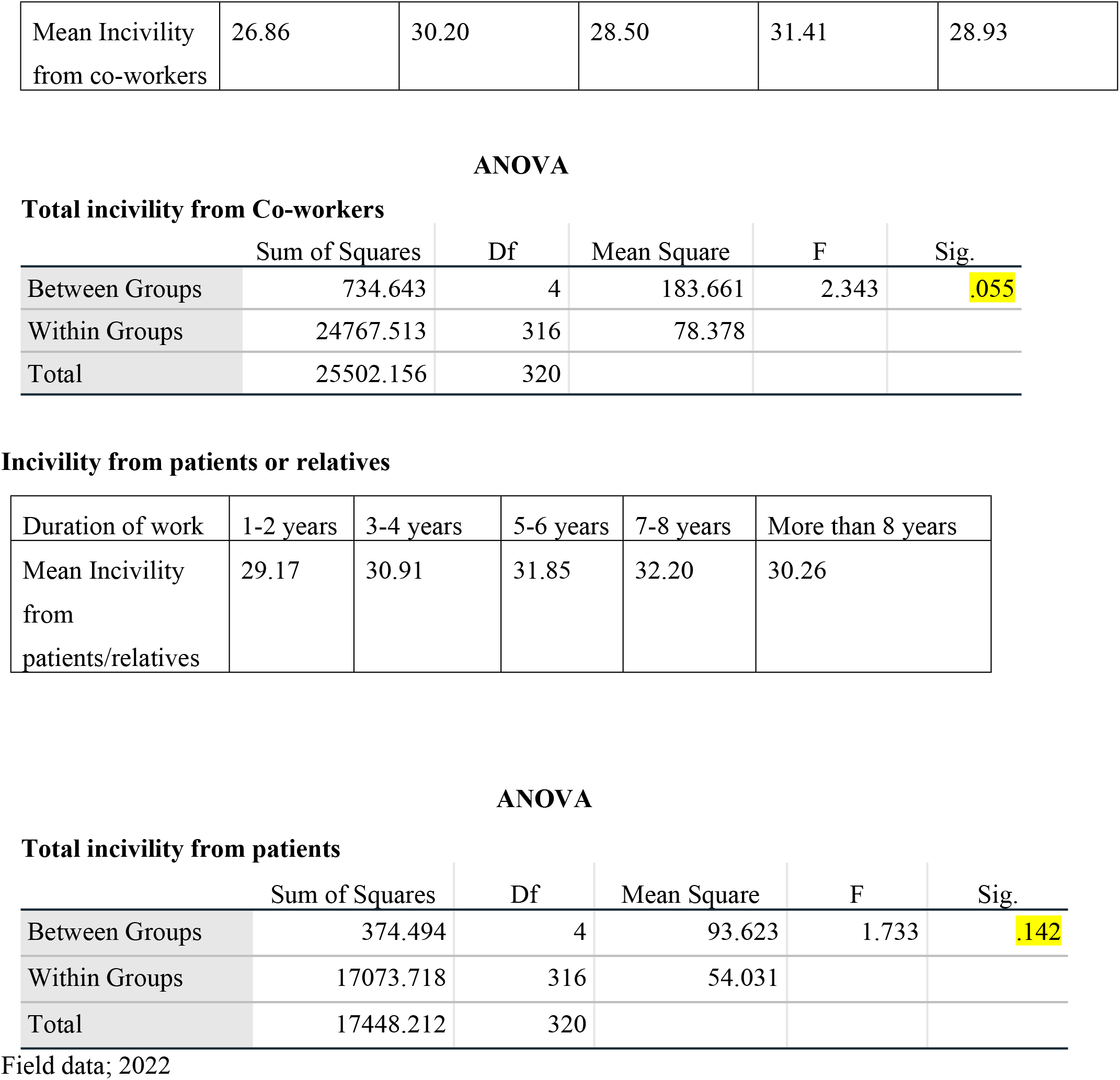
Effects of incivility on nursing and patient outcomes (safe work environment and intention to stay)

**Correlation**
| Spearman Rank Correlations | General incivility | Incivility from coworkers | Incivility from patients | Effects of incivility |
| --- | --- | --- | --- | --- |
| General incivility | 1 | .780 | .590 | -.276 |
| P-value | - | .000*** | .000*** | .000*** |

**General incivility**
| Duration of work | 1-2 years | 3-4 years | 5-6 years | 7-8 years | More than 8 years |
| --- | --- | --- | --- | --- | --- |
| Mean general Incivility | 25.72 | 28.00 | 27.26 | 29.14 | 28.25 |

**ANOVA** **Total General Incivility**
|  | Sum of Squares | Df | Mean Square | F | Sig. |
| --- | --- | --- | --- | --- | --- |
| Between Groups | 472.219 | 4 | 118.055 | 2.576 | .038 |
| Within Groups | 14484.024 | 316 | 45.836 |  |  |
| Total | 14956.243 | 320 |  |  |  |

**Incivility from nurses/co-workers**
| Duration of work | 1-2 years | 3-4 years | 5-6 years | 7-8 years | More than 8 years |
| --- | --- | --- | --- | --- | --- |
| Mean Incivility<br>from co-workers | 26.86 | 30.20 | 28.50 | 31.41 | 28.93 |

ANOVA Total incivility from Co-workers
|  | Sum of Squares | Df | Mean Square | F | Sig. |
| --- | --- | --- | --- | --- | --- |
| Between Groups | 734.643 | 4 | 183.661 | 2.343 | .055 |
| Within Groups | 24767.513 | 316 | 78.378 |  |  |
| Total | 25502.156 | 320 |  |  |  |

Incivility from patients or relatives
|  |  |  |  |  |  |
| --- | --- | --- | --- | --- | --- |
| Duration of work | 1-2 years | 3-4 years | 5-6 years | 7-8 years | More than 8 years |
| Mean Incivility<br>from<br>patients/relatives | 29.17 | 30.91 | 31.85 | 32.20 | 30.26 |

ANOVA Total incivility from patients
|  | Sum of Squares | Df | Mean Square | F | Sig. |
| --- | --- | --- | --- | --- | --- |
| Between Groups | 374.494 | 4 | 93.623 | 1.733 | .142 |
| Within Groups | 17073.718 | 316 | 54.031 |  |  |
| Total | 17448.212 | 320 |  |  |  |

Field data; 2022

From table 1.3 of the test statistics, there were mean differences between the duration of work and experiences of incivility from general, co-workers, and patients/relations. The highest mean incivility (n=29.14, 31.41 and 32.20) was experienced by participants with 7-8years duration of work experience from all the three sources of incivility (general, co-workers, and patients/relations). The mean values were 29.14, 31.41, and 32.20 respectively.

Again, from the test statistics above, there was a negative correlation between the experience of incivility (general, co-workers, and patients/relations) and effects on patients and nursing outcomes (safe work environment and intention to stay) with p-values of 0.000***, 0.000***, and 0.000*** respectively. Additionally, a p-value of 0.038 indicated statistical relevance between the number of years of employment and experiences with general incivility.

In conclusion, the prevalence of incivility in healthcare settings does not support a climate that is safe for nurses to provide safe care and the desire of nurses to stay. As a result, incivility in healthcare environments do not support the objective of fostering a secure working environment for nurses and patients.

## Discussion

### Prevalence and sources of incivility

The findings of this study revealed that incivility is very prevalent in the hospital with an overall average prevalence of 54.5%. These findings confirmed reports that incivility in healthcare settings is very endemic among nurses(3,4). For instance, according to reports from Western countries and Asia, the prevalence of incivility among nurses was 55% and 60%, respectively (5, 14).

In addition to the aforementioned results, the study findings showed that respondents’ average prevalence rates of incivility were higher and more diverse among the subscales. Under general, an overall average prevalent rate of 62.7% was reported with an average highest prevalence rate of 71.6% for hostile climate subscale. Also, displaced frustration subscale under patients/relations recorded the highest average prevalent rate of 78.9%. In essence, the prevalence of incivility in healthcare settings does not promote respect as a workplace norm among nurses. This results in ineffective teamwork among healthcare team members which puts patient safety at risk (10, 11).

Furthermore, the findings showed that incivility was perpetrated frequently by employees who were not either fellow nurses, direct supervisors, and physicians/doctors but general staff (general incivility) and patients/relations. The study findings also revealed that the least incivility was also sometimes, most of the time, and all of the time perpetrated by direct nurse supervisors. For instance, the study findings revealed that nurses in the hospital experienced incivility sometimes, most of the time, and all the time more from patients/relations and general. Again, the study findings also reported that nurses experienced incivility sometimes, most of the time, and all the time least from their direct supervisors. These findings corroborate with a study by Alquwez (10) which reported that nurses experience incivility most from patients/visitors and least incivility from their supervisors. The reasons being that nurses accept acts of incivility as part of professional and dysfunctional work environment norms (15). As a result, nurses especially, inexperienced nurses, are usually hesitant to give negative responses about incivility experienced from supervisors, nurse managers for fear of revenge and victimisation. Again, Layne and colleagues reported that nurses experienced higher incidence of incivility from patients/family most of the time than other sources such as physicians, coworkers, other employees and supervisors. This is evident in the study findings which showed high incivility rate among general and patients/relations. Furthermore, as reported by Gardner (16), new nurses are always hesitant to give negative responses in a questionnaire measuring experiences of incivility at the workplace.

In conclusion, nurses experienced high prevalence of incivility from patients/relations and general sources. These study findings corroborated with other findings in literature. In addition, the prevalence of incivility from these sources were highest under the displaced frustration subscale under patients/relations and hostile climate subscale under general incivility sources respectively. This implies that, though nurses could experience incivility from general, he/she could experience highest prevalence from hostile climate or inappropriate jokes subscales of the general incivility as well as other subscales from direct supervisors, patients/relations, physicians/doctors, and co-worker’s incivilities respectively.

### Effects of incivility on patient and nursing outcomes (safe work environment and intention to stay)

The study also sought to examine whether there are any associated effects of the experiences of incivility by nurses on patients and nursing outcomes. The effects were examined through the existence of safe work environment and intention to stay or leave by nurses.

The findings of this study showed that there is a negative correlation between the experiences of general, patients/relations, co-worker’s incivilities, and patients and nursing outcomes (safe work environment, intention to stay) with p-values of 0.000***, 0.000*** and 0.000*** respectively. This implies that the experiences of incivility from general, patients/relations, and co-worker’s sources promote unsafe work environments. Again, nurses who experienced incivility from any of these sources do not have the zeal to stay and work in such settings thereby increasing the intent to leave than stay. These findings are consistent with a study by Alquwez (10), that concluded that nurses’ experiences of incivility were associated with negative patient safety competence and quality of nursing care. Furthermore, Phillips and colleagues (17), reported that the presence of incivility in the workplace and among nurses interfere with patient safety. Acts of incivility experienced by junior nurses deter them from consulting experienced ones in difficult clinical situations for fear of uncivil behaviours thereby putting patient safety at risk. Additionally, nurses who perceive the work environment negatively due to acts of low intensity disruptive behaviours (incivility) leave their departments or the hospital, resulting in high staff turnover rates. Also, a study by Shoorideh and colleagues (18), showed that the presence of incivility among nurses in healthcare settings is a predictor of low job satisfaction and performance which leads to low quality nursing care and unsafe care. Again, a study by Gardner (16), reported that incivility among healthcare personnel is associated with unsafe work environment and negative nursing outcomes (decreased job satisfaction, increased intention to leave). More so, Phillips and colleagues (17), concluded that about one million patients die worldwide due to unsafe care resulting from incivility among healthcare professionals in healthcare settings.

## Limitation of the study

A cross-sectional study design was used that inhibits causal relationships between variables. In addition, the sampling was done from one specific occupation in one tertiary hospital in one geographic area in the country thereby limiting the generalisability of the findings. Despite these limitations in the generalisability of the findings, healthcare professionals and policy makers would find these findings useful in creating awareness about the phenomenon in healthcare settings.

## Conclusion

The study findings revealed that incivility is very prevalent in healthcare settings. The prevalence varies considerably among the nursing incivility subscales. For instance, though the overall average incivility prevalence in this study was 54.5%, the highest prevalence was reported from displaced frustration subscale of 78.9% under patients/relations, followed by the least prevalent of 39.9% from direct supervisors. The study findings also revealed that nurses experienced incivility frequently from general staff and patients/relations.

In addition, the study findings revealed that, there is a negative association between the experiences of incivility by nurses from general, patients/relations, co-workers and patients and nursing outcomes (safe work environment and intention to stay or leave by nurses). The study findings corroborated with many studies which investigated the perilous effects of incivility.

## Data Availability

All relevant data are within the manuscript and its Supporting Information files.

## Acknowledgement

The authors are grateful to the respondents for the support and cooperation they obtained during the data collection at the hospital. Our appreciation goes to the management of the hospital for giving administrative approval for the conduct of the study.

## References

1. Cook C. Climate change and health: Nurses as drivers of climate action. Interdisciplinary Journal of Partnership Studies. 2018 Feb 26;5(1):8-.

2. Simpson KR. Incivility, bullying, and workplace violence: New recommendations for nurses and their employers from the American Nurses Association. MCN: The American Journal of Maternal/Child Nursing. 2016 Jan 1;41(1):68.

3. Amoo SA, Menlah A, Garti I, Appiah EO. Bullying in the clinical setting: Lived experiences of nursing students in the Central Region of Ghana. PLoS one. 2021 Sep 23;16(9):e0257620.

4. Phillips JM, Stalter AM. Systems thinking for managing covid-19 in health care systems: Seven key messages. The Journal of Continuing Education in Nursing. 2020 Sep 1;51(9):402–11.

5. Conner Black A. Promoting Civility in Healthcare Settings. International Journal of Childbirth Education. 2019 Apr 1;34(2).

6. Hopkinson SG, Dickinson CM, Dumayas JY, Jarzombek SL, Blackman VS. A multi-center study of horizontal violence in United States military nursing. Nurse education in practice. 2020 Aug 1;47:102838.

7. Houck NM, Colbert AM. Patient safety and workplace bullying: an integrative review. Journal of nursing care quality. 2017 Apr 1;32(2):164–71.

8. Kenawy AS, Kett V. The impact of electronic prescription on reducing medication errors in an Egyptian outpatient clinic. International journal of medical informatics. 2019 Jul 1;127:80–7.

9. Bloomfield, J. and Fisher, M.J., 2019. Quantitative research design. Journal of the Australasian Rehabilitation Nurses Association, 22(2), pp.27–30.

10. Alquwez N. Examining the influence of workplace incivility on nurses’ patient safety competence. Journal of Nursing Scholarship. 2020 May;52(3):292–300.

11. Armstrong NE. A quality improvement project measuring the effect of an evidence-based civility training program on nursing workplace incivility in a rural hospital using quantitative methods. Online Journal of Rural Nursing and Health Care. 2017 Mar 6;17(1):100–37.

12. Layne DM, Anderson E, Henderson S. Examining the presence and sources of incivility within nursing. Journal of Nursing Management. 2019 Oct;27(7):1505–11.

13. Guidroz AM, Burnfield-Geimer JL, Clark O, Schwetschenau HM, Jex SM. The nursing incivility scale: Development and validation of an occupation-specific measure. Journal of nursing measurement. 2010 Dec 1;18(3):176–200.

14. Ma C, Meng D, Shi Y, Xie F, Wang J, Dong X, Liu J, Cang S, Sun T. Impact of workplace incivility in hospitals on the work ability, career expectations and job performance of Chinese nurses: a cross-sectional survey. BMJ open. 2018 Dec 1;8(12):e021874.

15. Crawford CL, Chu F, Judson LH, Cuenca E, Jadalla AA, Tze-Polo L, Kawar LN, Runnels C, Garvida Jr R. An integrative review of nurse-to-nurse incivility, hostility, and workplace violence: a GPS for nurse leaders. Nursing administration quarterly. 2019 Apr 1;43(2):138–56.

16. Gardner J. Incivility among nurses, the influence of structural empowerment: A systematic review (Doctoral dissertation, Walden University).

17. Phillips JM, Stalter AM, Winegardner S, Wiggs C, Jauch A. Systems thinking and incivility in nursing practice: An integrative review. InNursing forum 2018 Jul (Vol. 53, No. 3, pp. 286–298).

18. Shoorideh FA, Moosavi S, Balouchi A. Incivility toward nurses: a systematic review and meta-analysis. Journal of Medical Ethics and History of Medicine. 2021;14.

